# Machine Learning-Based Prediction of Free IgE Concentration in Allergic Rhinitis Patients Treated with Allergen Immunotherapy and Omalizumab

**DOI:** 10.1101/2023.09.29.23296326

**Authors:** Kazeem B. Olanrewaju, Laura Marthe Emilie Ngansop Djampou

## Abstract

Free immunoglobulin E (IgE) concentration is a key biomarker for allergic diseases. Prediction of free IgE concentration can help clinicians diagnose and monitor allergic diseases more effectively. In this study, we used machine learning to predict free IgE concentration in the blood serum of patients with allergic rhinitis who received allergen immunotherapy co-administered with omalizumab. The predictors for free IgE concentration were the number of visits for treatment and baseline checking, and treatment groups (1) omalizumab/ragweed, (2) omalizumab/placebo, (3) placebo/ragweed, and (4) placebo/placebo. Several machine learning algorithms (MLA) were trained with the immunotherapy dataset imported from Immune Tolerance Network (ITN) TrialShare into the Orange data mining platform. The decision tree algorithm model amidst the list of MLAs trained and tested was the best performing model for predicting free IgE concentration, with an R-squared of about 0.6. This study demonstrates that machine learning can be used to predict free IgE concentration with high accuracy. This prediction model could be used to help clinicians diagnose and monitor allergic diseases more effectively.

## Background and Significance

Immunoglobulin E (IgE) is an antibody that plays a key role in the development of allergic diseases. When a person with allergies is exposed to an allergen, their body produces IgE antibodies that bind to the allergen^1, 2^. This binding triggers the release of histamine and other inflammatory mediators, which cause the symptoms of an allergic reaction^3^. Free IgE concentration in the blood is a measure of the amount of IgE antibodies that are not bound to allergens. Free IgE concentration is a key biomarker for allergic diseases, and it can be used to diagnose and monitor these diseases^4, 5^. Allergen immunotherapy (AIT) is a treatment that involves exposing a person to gradually increasing doses of an allergen to desensitize them to the allergen. Omalizumab is a monoclonal antibody that targets IgE^6^. It is used to treat severe allergic asthma and other allergic diseases. Prediction of free IgE concentration can help clinicians diagnose and monitor allergic diseases more effectively. For example, if a patient has a high free IgE concentration, it is more likely that they have an allergy. Additionally, monitoring free IgE concentration over time can help clinicians track the effectiveness of AIT and omalizumab treatment^7-9^. Machine learning is a type of artificial intelligence that can be used to learn from data and make predictions. Machine learning algorithms are gradually gaining ground as tools that can be very effective at predicting free IgE concentration and other immunological biomarkers in patients with allergic diseases^10-12^. In this study, we are going to use a machine learning algorithm to predict free IgE in allergic rhinitis patients treated with allergen immunotherapy and omalizumab^5, 6, 9, 13^. The development of machine learningbased prediction models for free IgE concentration in patients with allergic rhinitis treated with AIT and omalizumab would be a significant advance in the field of allergy management. These models could be used to help clinicians diagnose and monitor allergic rhinitis more effectively, and to track the effectiveness of AIT and omalizumab treatment. Potential applications of machine learning-based prediction models for free IgE concentration in patients with allergic rhinitis treated with AIT and omalizumab:

### Diagnosis

Machine learning models could be used to develop diagnostic algorithms for allergic rhinitis. These algorithms could be used to identify patients with allergic rhinitis based on their free IgE concentration and other factors^14-17^.

### Monitoring

Machine learning models could be used to monitor the effectiveness of AIT and omalizumab treatment in patients with allergic rhinitis. This would allow clinicians to adjust the treatment plan as needed to ensure that the patient is receiving the most effective treatment possible^15, 18, 19^.

### Personalized treatment

Machine learning models could be used to develop personalized treatment plans for patients with allergic rhinitis. These treatment plans could be based on the patient’s free IgE concentration, other factors, and the patient’s preferences^20-22^. Overall, the development of machine learning-based prediction models for free IgE concentration in patients with allergic rhinitis treated with AIT and omalizumab has the potential to significantly improve the management of allergic rhinitis.

## Research Methodology

### Dataset Extraction from Immune Tolerance Network/TrialShare

The Immune Tolerance Network ITN/TrialShare^23^ repository contains data from clinical trials of immunotherapy, including both mechanistic and raw clinical datasets. The ITN TrialShare dataset will be used as the primary dataset for this study, as it is expected to be the most comprehensive and up-to-date dataset available. To extract the immunotherapy biomarker dataset^9, 13^ from the ITN TrialShare repository, the following steps will be followed:

1. Access the data repository by logging into the TrialShare link on the Immune Tolerance Network (ITN) page.
2. Filter the studies of interest by selecting the therapeutic area (allergy), study type (clinical trial), age group (all adults), phase (all phases), and condition (allergic rhinitis).
3. Select the appropriate study or studies by clicking the “GO TO STUDY” button.
4. On the study page, export the dataset as an Excel file (xlsx).

The dataset will then be uploaded into Orange Data Mining Platform^24^ (the Machine Learning Software Platform (python-based) for this study) as CSV files to undergo the various steps leading to the development of the machine learning algorithm model for Free Ige Concentration prediction.

### Dataset Preprocessing

The first step in this process is to visualize and preprocess the data to address a range of issues that may affect the learning of the data by the several machine learning algorithms considered before selecting the best performing algorithm for the free IgE concentration predictive model. Preprocessing of the dataset will involve the following steps:

1. Data cleaning: This step involves addressing inconsistency, duplicity, noise, and missing data.
2. Data integration: This step involves combining relevant datasets from the mechanistic and the clinical datasets.
3. Data reduction: This step involves reducing data attributes and dimensionality to features with significant impact (high information gain) on the target variable (s).
4. Data transformation: This step involves creating a function that can map old values into a new set of values through smoothing functions (Fourier transform), feature construction, aggregation, and normalization such that each old value is identified with a new value.
5. Data discretization: This step involves applying interval or conceptual labels to datasets such as age groups and immunotherapy patients’ weekly injection duration.

Once the data has been preprocessed, it will be ready for use in the machine learning algorithm model development process.

### Machine Learning Algorithm Training and Testing

The preprocessed dataset will advance to the next stage within the Orange data mining platform, which focuses on machine learning and evaluation, specifically the prediction phase. During this step, the identification and categorization of the dataset attributes into predictors and target variables will be done. Free IgE concentration will be designated as the target variable while the treatment group ((1) omalizumab/ragweed, (2) omalizumab/placebo, (3) placebo/ragweed, and (4) placebo/placebo), V_visit_common (weekly visit no treatment administered), and V_Visit_num (treatment IgE assessment weekly visit with treatment administered). Other attributes were not considered in the predictors list because they are attributes meant for participant identification. These attributes are treated as meta-data or simply ignored in the machine learning algorithm procedures. Typically, machine learning employs a single variable as the target, but occasionally, multiple variables can be designated as targets simultaneously. This machine learning approach, where there is a well-defined target variable, is known as supervised machine learning. Among the supervised machine learning algorithms under consideration are k-nearest neighbors (kNN), decision trees, linear regression, random forests, artificial neural networks (ANN), gradient boosting, and support vector machines.

The training of the machine learning algorithm will consist of using 70-80% of the dataset for training and reserving 20-30% for testing. During this process, the algorithm was tuned (sensitivity analysis) by employing techniques such as cross-validation with various fold numbers, bootstrapping for random forest algorithms, and adjusting algorithm parameters, such as the number of nearest neighbors in kNN or modifying neurons and activation methods in ANN. After the training phase is completed, the testing of the machine learning algorithm on the dataset immediately follows. This machine learning process within the Orange Data Mining platform will involve a sequence of widgets encompassing data manipulation, transformation, visualization, modeling, and evaluation. These widgets are Python-based tools provided by the Orange Data Mining platform.

Initial datasets have about 5.2 % missing values which are completely from the free IgE concentration. The missing data were addressed during the preprocessing stage in three ways. These include filling the missing free IgE concentration values with the average/most frequent values, replacing them with random values, removing rows with missing values, and replacing them with predicted values from the bestperforming MLA model. Thus, the machine learning algorithm with the best-performing metrics, including coefficient of determination (R^2^), mean squared error (MSE), root mean squared error (RMSE), and mean absolute error (MAE) will be chosen as the Machine Learning Algorithm Model for predicting the Free IgE concentration. Amidst the list of MLA performance metrics, the coefficient of determination (R^2^) will be the top determinant metric in making the best selection.

## Results and Discussion

### Different scenarios for addressing missing values and the performance metric results

#### Scenario 1: imputing missing values with the average/most frequent value

The missing values in the datasets were addressed by imputing missing free IgE values with the average/most frequent with cross-validation of 10 folds and 80 % sampled dataset for MLA training and 20% dataset to test the MLA model for free IgE concentration prediction. In addition, the parameter for each MLA considered was tuned to achieve the optimum performance metric. The resultant performance metric for this approach can be seen in the Test and Score widget window below. Figure 1 below shows the metric performance measured for MLA trained and tested in this scenario. Thus, the decision tree, the best performing MLA model, has its optimum parameter at 29 minimum number of instances in leaves with split subsets not smaller than 80. The worst-performing MLA model, linear regression, has its optimum parameter at alpha 30 for the ridge regression as depicted in Figure 2.

**Figure 1.**
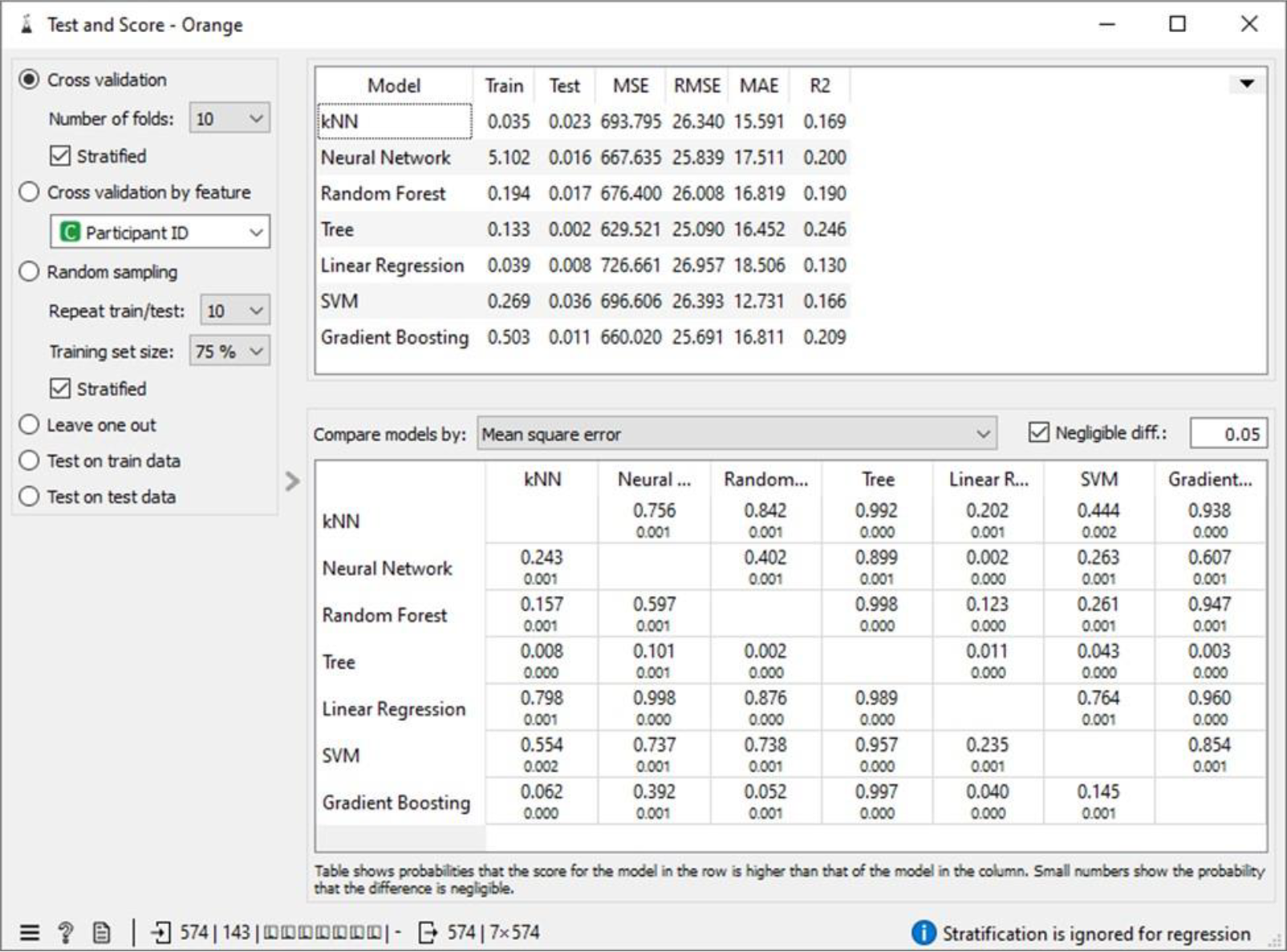
Test and Score showing the model evaluation metric for the MLAs under scenario 1

**Figure 2.**
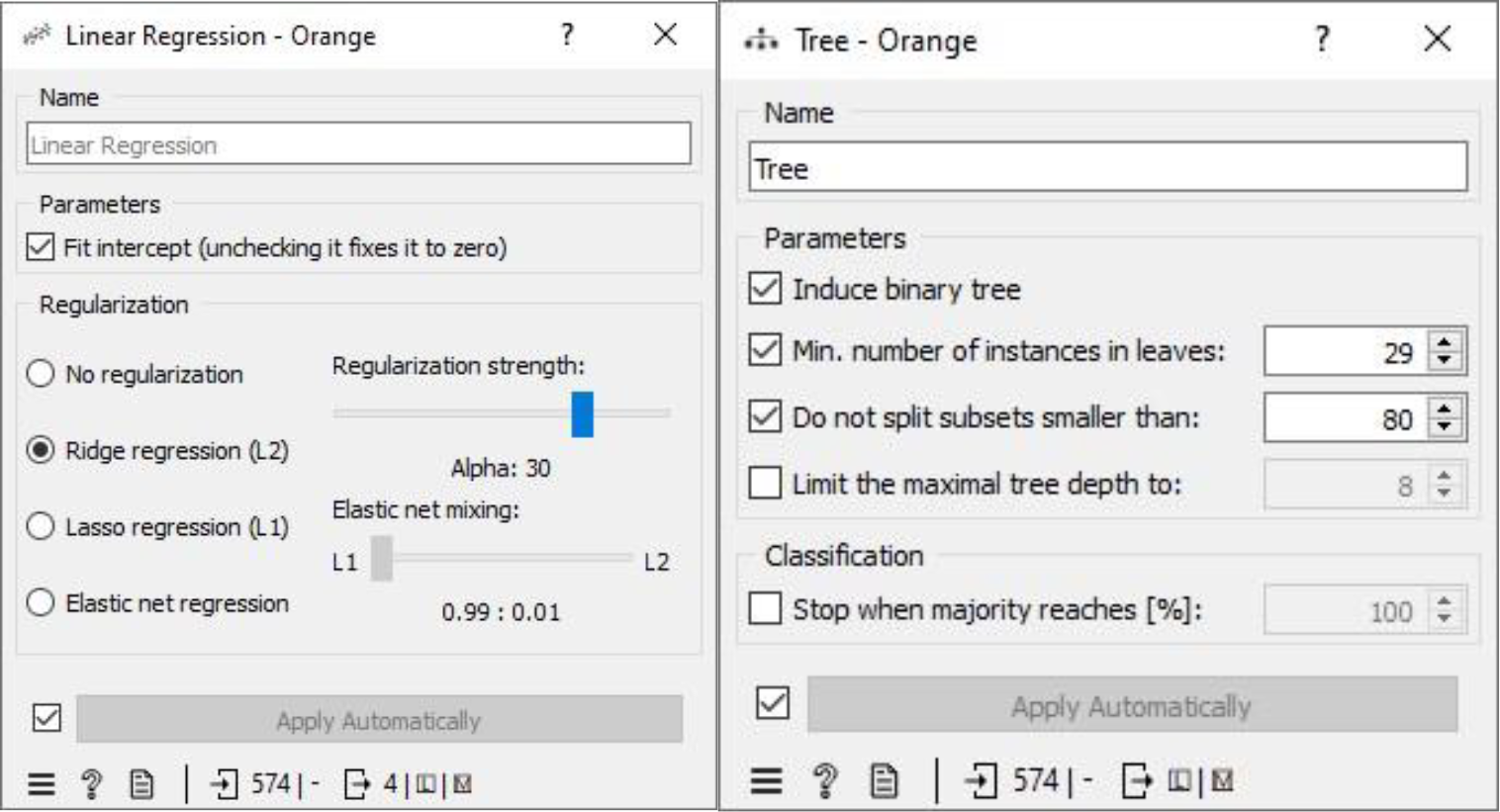
Linear regression and Decision Tree parameter windows under scenario 1

#### Scenario 2: Replacing missing values with random value

Missing values in the dataset were replaced with random values. Then, the dataset was split into 10 folds, and 80% of the data was used to train the machine learning algorithms. The remaining 20% of the data was used to test the algorithms and to tune their parameters. The decision tree algorithm was the bestperforming algorithm, with the lowest errors measured relative to other algorithms. Its optimum parameter was 29 minimum number of instances in leaves with split subsets not smaller than 120. The worst performing algorithm was support vector machine (SVM), with its optimum parameters at Cost (c) of 24.00 and regression loss epsilon (ε) equals 4.10 with RBF kernel.

#### Scenario 3: Removing rows with missing values option

Missing values in the dataset were handled by applying 10-fold cross-validation. We allocated 80% of the dataset for training machine learning algorithms (MLA) and reserved 20% for testing the MLA models used to predict free IgE concentration. This process led to a reduction in the dataset size from 717 instances to 418. The parameters of each MLA were fine-tuned to achieve optimal performance metrics. The results of this approach, including the performance metrics, are available in the Test and Score widget window. Figure 5 displays the performance metrics for the MLAs trained and tested in this context. Among the considered MLAs, the neural network emerged as the best-performing model, as illustrated in Figure 6. Its optimal configuration included three hidden layers with 50, 100, and 50 neurons, utilizing ReLu activation, and a regularization parameter (α) set to 0.05. The algorithm was trained and tested for a maximum of 500 iterations. Conversely, the least effective MLA model among those considered was linear regression, specifically the ridge regression variant, which achieved its best performance with an alpha value of 30, as indicated in Figure 6.

**Figure 3.**
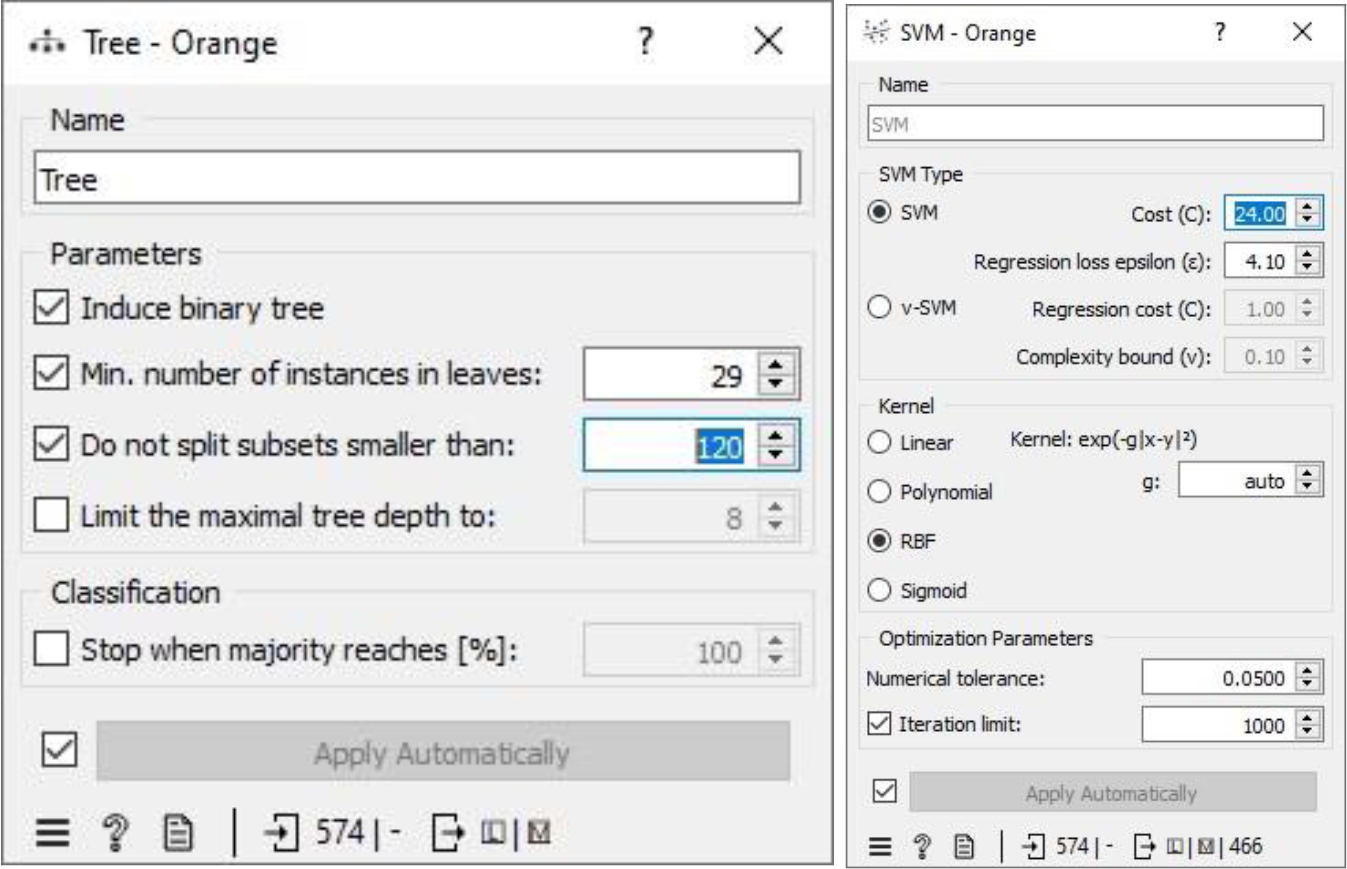
Decision Tree and Support Vector Machine parameter windows under scenario 2

**Figure 4.**
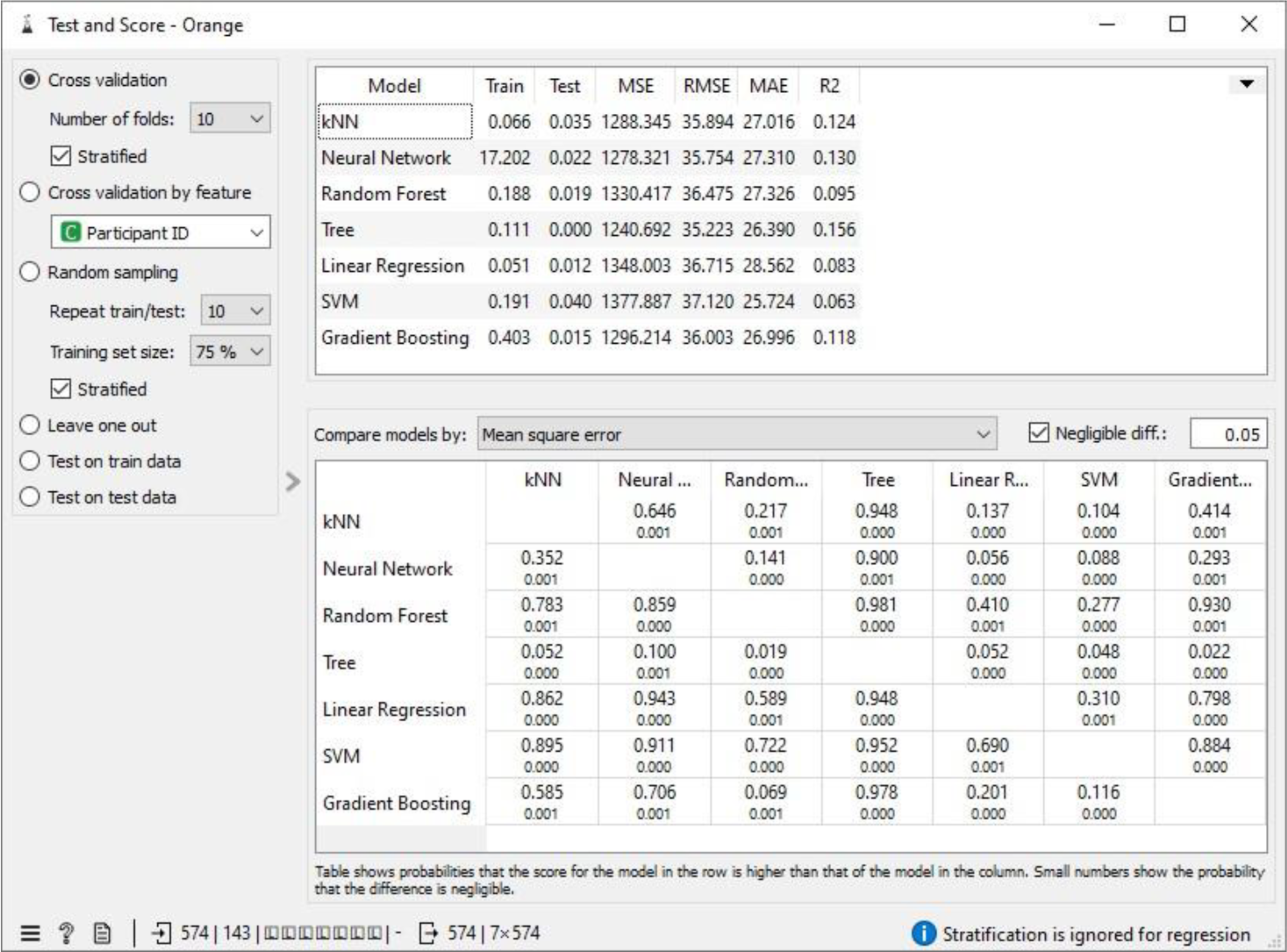
Test and Score showing the model evaluation metric for the MLAs under scenario 2

**Figure 5.**
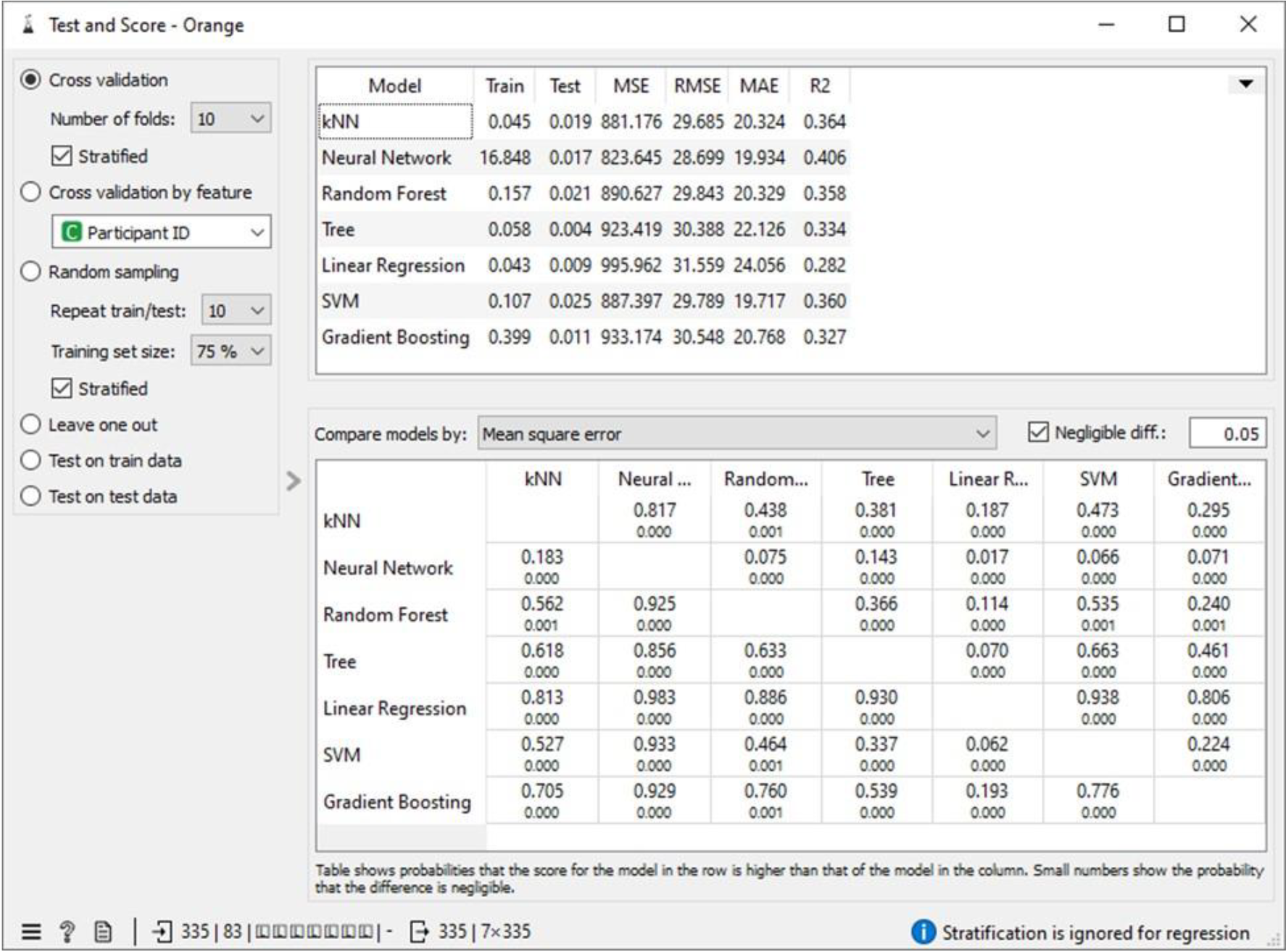
Test and Score showing the model evaluation metric for the MLAs under scenario 3

**Figure 6.**
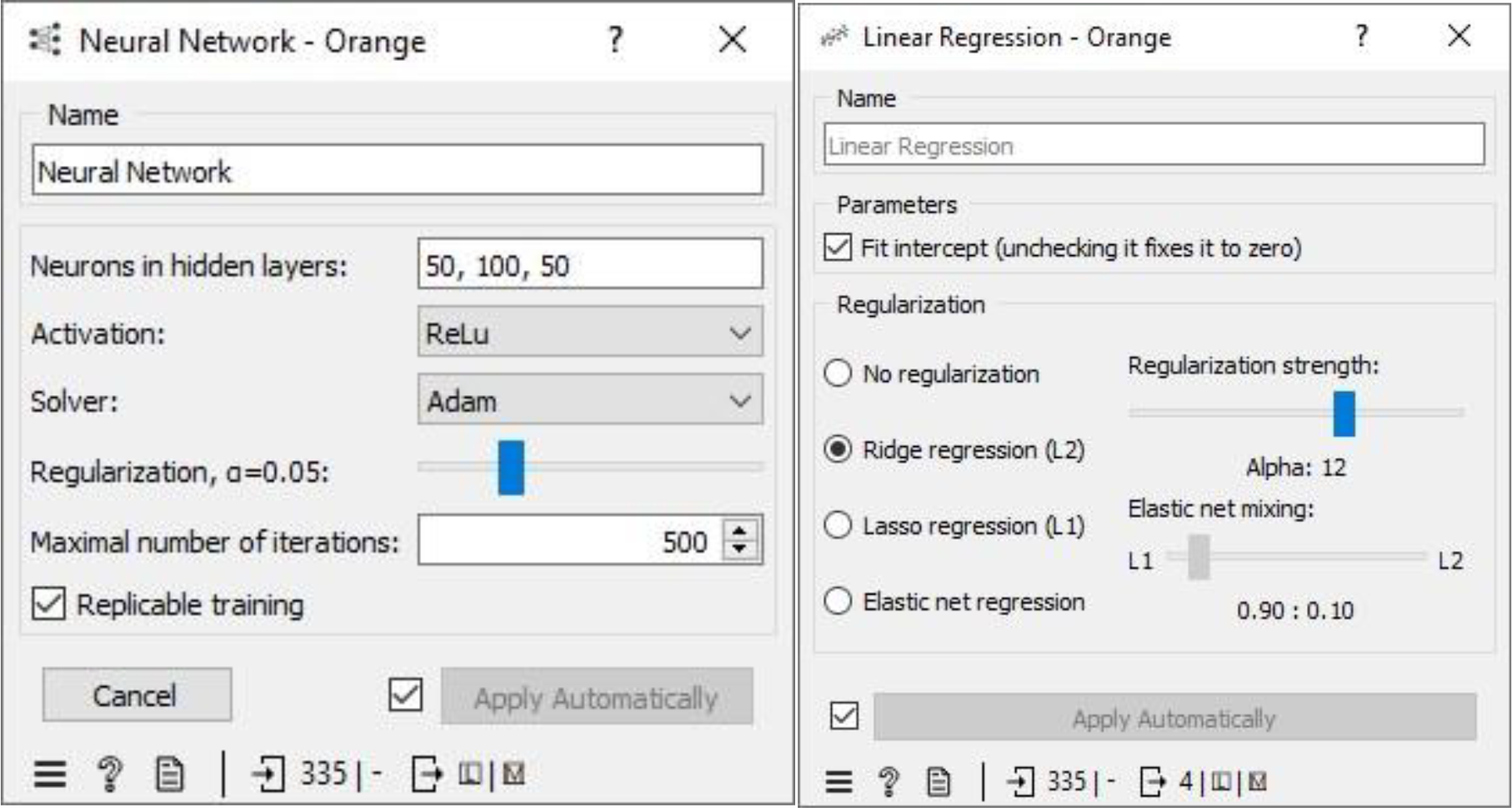
Neural Network and Linear Regression parameter windows under scenario 3

#### Scenario 4: Replacing with missing values with machine-learning-algorithm-generated data

To handle missing values in the datasets, we employed a method of imputation by replacing the missing free IgE values with predictions made by an MLA (Machine Learning Algorithm) specifically designed for this task. These predictions were generated using measured features that corresponded to the missing free IgE concentrations, serving as predictor variables. Notably, the predictive model used for this purpose was a neural network MLA. The machine pipeline for this scenario can be seen below in Figure 7. Our approach involved training the MLAs using 80% of the dataset with a 10-fold cross-validation strategy, while the remaining 20% of the dataset was reserved for testing. This division allowed us to identify the most proficient MLA for predicting free IgE concentrations. Furthermore, we meticulously fine-tuned the parameters of each MLA to optimize various performance metrics, including coefficient determination (R2), mean absolute error (MAE), root mean square error (RMSE), and mean square error. The performance metrics resulting from this method can be observed in the Test and Score widget window, and Figure 8 visually represents the performance metrics for the MLAs trained and tested within this context. The decision tree MLA emerged as the top-performing model, as depicted in Figure 8. Its optimal configuration featured a minimum of 29 instances in leaves, with split subsets not smaller than 120. The neural network, which ranked as the next best-performing model, was configured with three hidden layers comprising 50, 100, and 50 neurons, utilizing ReLu activation and a regularization parameter (α) set to 0.05. Additionally, the maximum number of iterations employed during the algorithm’s training and testing phases was limited to 500. Conversely, the least effective MLA model among those considered was linear regression, particularly the ridge regression variant, which demonstrated its best performance with an alpha value of 12, as displayed in Figure 9.

**Figure 7.**
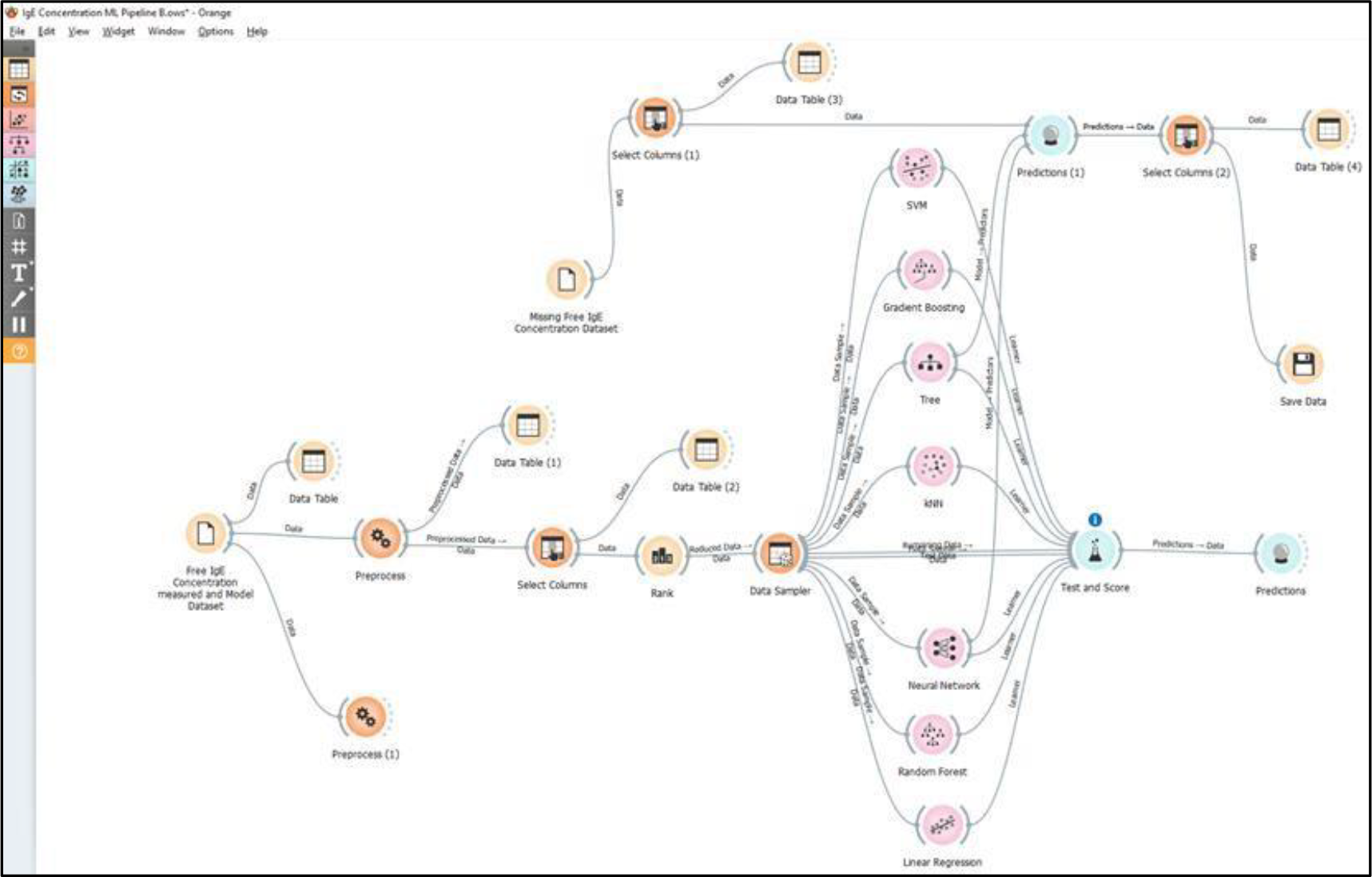
Machine Learning Pipeline for the Prediction of Free Immunoglobin E (IgE)

**Figure 8.**
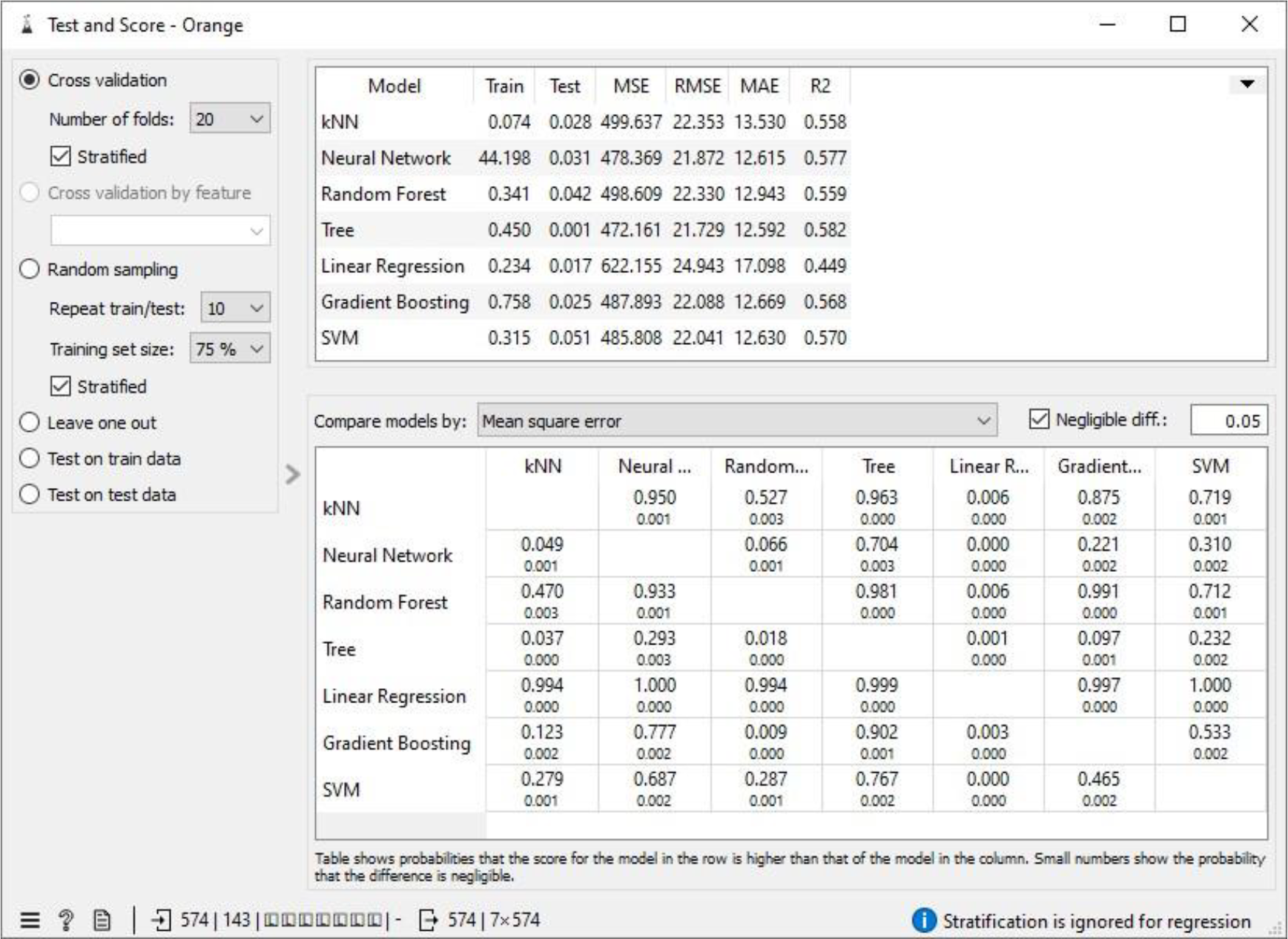
Test and Score showing the model evaluation metric for the MLAs under scenario 4

**Figure 9.**
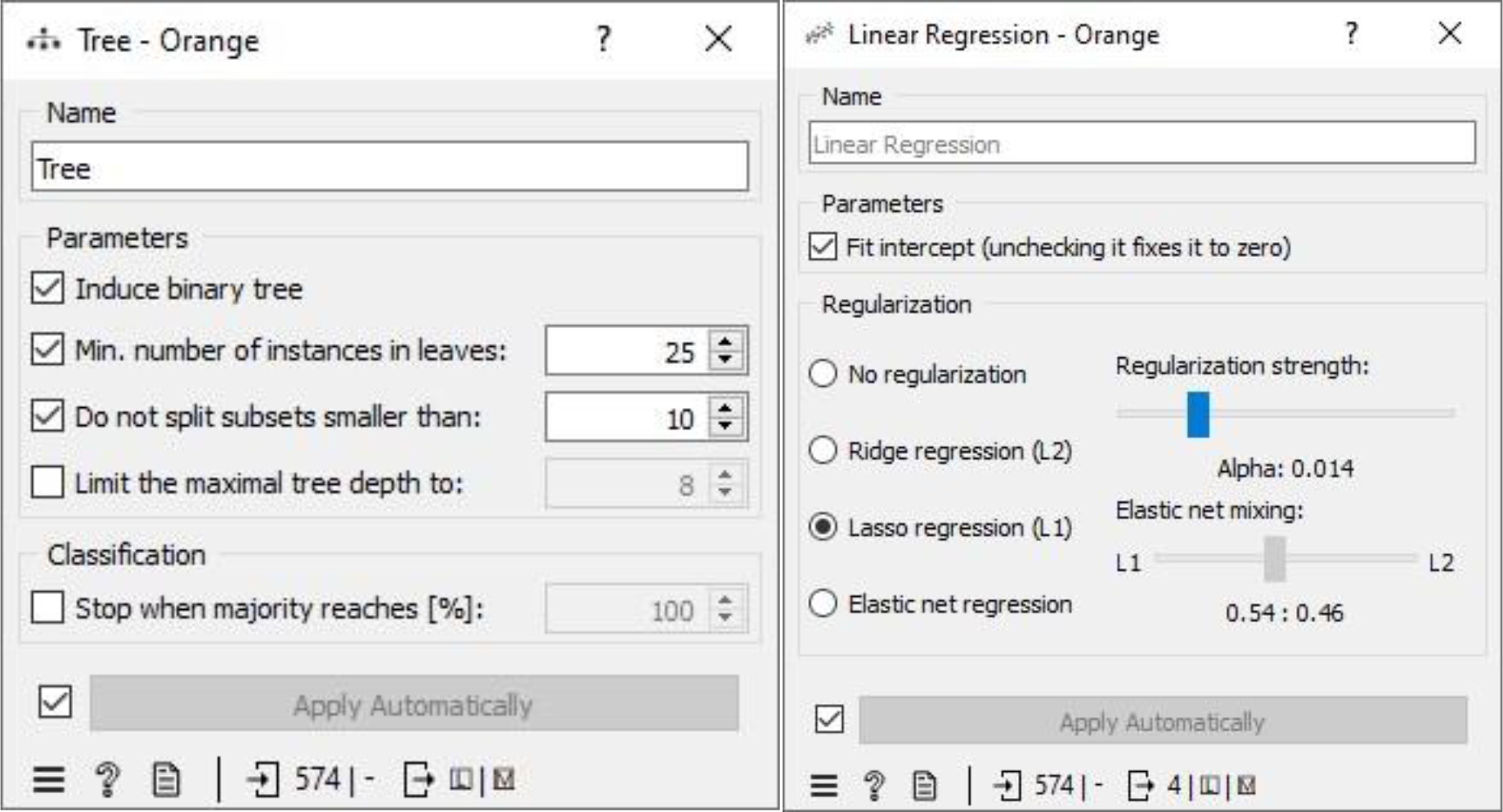
Neural Network and Linear Regression parameter windows under scenario 4

Scenario 4 yields the most favorable performance evaluation results when predicting free IgE concentration based on three crucial predictor variables: treatment group (TRT_Grp), weekly visits without treatment administered (V_visit_num), and weekly visits with treatment administered (V_visit_common). The coefficient of determination (R2) for scenario 4 which is 0.582 is significant for a dataset involving humans. Additionally, we conducted an assessment of information gain for these three predictor variables, which measures their impact on predicting the free IgE concentration. This analysis revealed that the treatment groups (TRT_Grp) exert a substantial influence on predicting free IgE concentration. In contrast, the impact of weekly visits, both with and without treatment administered, was found to be negligible in terms of predicting free IgE concentration. These findings align with the insights provided in the Rank widget of the Orange Data Mining platform, as illustrated in Figure 10. It reinforces the statement made on the Free IgE Dataset Properties page of the ITN TrialShare, which indicates that the dataset lacks a clear relationship with specific visits.

**Figure 10.**
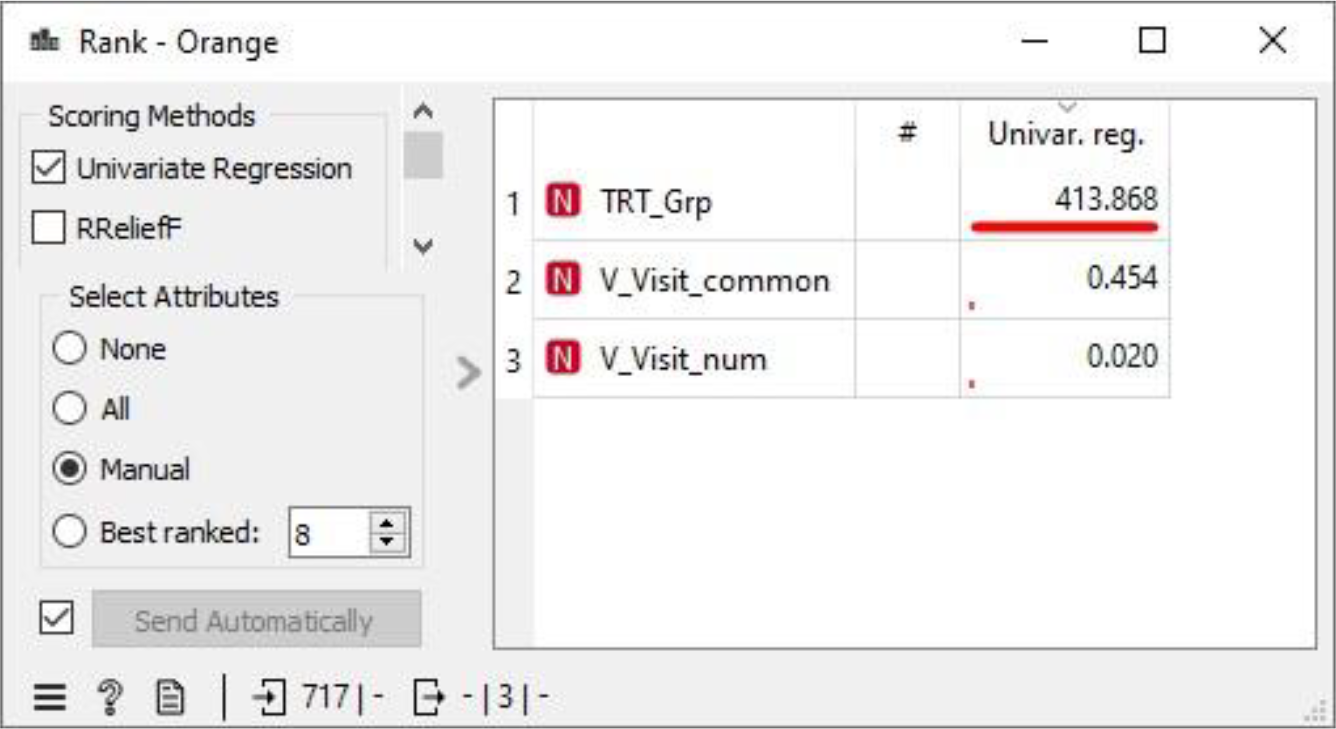
Impact of the three attributes in predicting the Free IgE concentration

Table 1 shows assessment metrics for all the machine learning algorithms (MLAs) trained and tested with the Free IgE concentration dataset. The neural network algorithm spent almost 45 seconds in learning the dataset and 0.031 seconds in testing while 0.074 seconds was used to train the kNN model and 0.028 seconds was used to the test data. The best performing MLA model in Table 1 is the decision tree with coefficient of determination, R^2^ = 0.582; and the lowest errors measured relative to other MLA models. These errors include mean square error (MSE =472.161), root mean square error (RMSE= 21.79), and mean absolute error (MAE=12.592).

**Table 1.** Showing the metric for the scenario 4.

|  | **Scenario 4** |  |  |  |  |  |
| --- | --- | --- | --- | --- | --- | --- |
| **MLA Model** | **Train time (s)** | **Test time (s)** | **MSE** | **RMSE** | **MAE** | **R<sup>2</sup>** |
| **kNN** | 0.074 | 0.028 | 499.637 | 22.353 | 13.530 | 0.558 |
| **Neural Network** | 44.198 | 0.031 | 478.369 | 21.872 | 12.615 | 0.577 |
| **Random Forest** | 0.341 | 0.042 | 498.609 | 22.330 | 12.943 | 0.559 |
| **Tree** | **0.450** | **0.450** | **472.161** | **21.729** | **12.592** | **0.582** |
| **Linear regression** | 0.234 | 0.017 | 622.155 | 24.943 | 17.098 | 0.449 |
| **Gradient Boosting** | 0.785 | 0.025 | 487.893 | 22.088 | 12.699 | 0.568 |
| **SVM** | 0.315 | 0.051 | 485.808 | 22.041 | 12.630 | 0.570 |

## Conclusion

This study aims to develop a machine-learning model for predicting free IgE concentration in patients with allergic rhinitis treated with AIT and omalizumab. The dataset was extracted from Immune Tolerance/TrialShare, which contains data from clinical trials of immunotherapy. The datasets were preprocessed to address inconsistency, duplicity, noise, and missing data. Several machine learning algorithms were trained and tested on the preprocessed dataset. The algorithm with the best-performing metrics was chosen as the Machine Learning Algorithm Model for predicting the Free IgE concentration. Potential applications of the machine learning model include diagnosis, monitoring, and personalized treatment of allergic rhinitis. Four scenarios which include replacing the missing values with average/most frequent, random values, removing instances with missing free IgE concentration, and replacing missing IgE concentration with MLA predicted values were adopted to address the missing free IgE concentration in the dataset. Scenario 4 gave the best performance for the prediction of free IgE concentration based on the three predictor variables. The coefficient of determination (R2) for scenario 4 is 0.582, which is significant for a dataset involving humans. The treatment group has a major impact on the free IgE concentration prediction, while weekly visits with and with no treatment administered have negligible impact on predicting the free IgE concentration. The decision tree algorithm is the bestperforming algorithm with the lowest errors measured relative to other MLA models.

## Data Availability

All data produced in the present study are available upon reasonable request to the authors.

